# Transcatheter Versus Surgical Aortic Valve Replacement in Medicare Beneficiaries with Bicuspid Aortic Stenosis (TREAT-BICUSPID): Statistical Analysis Plan

**DOI:** 10.1101/2025.03.09.25323634

**Authors:** Parth N. Patel, Olivia L. Hulme, Huaying Dong, Yang Song, David J. Cohen, Robert W. Yeh, Dhaval Kolte

## Abstract

This document provides the full statistical analysis plan (SAP) for the TRanscatheter VErsus Surgical Aortic Valve ReplacemenT in Medicare Beneficiaries with Bicuspid Aortic Stenosis (TREAT-BICUSPID) study. TREAT-BICUSPID is a retrospective, population-based cohort study of Medicare beneficiaries with bicuspid aortic stenosis (AS) undergoing aortic valve replacement (AVR) from 2012 to 2022. The study was designed to evaluate trends in relative use of transcatheter aortic valve replacement (TAVR) and surgical aortic valve replacement (SAVR) in this population over time, and compare 1-year outcomes of isolated TAVR and SAVR using an instrumental variable approach. The pre-specified SAP detailed in this document was posted on medRxiv prior to analysis of the primary outcomes.

## Overview

Multiple landmark clinical trials over the last 15 years comparing transcatheter aortic valve replacement (TAVR) and surgical aortic valve replacement (SAVR) across all levels of surgical risk have established TAVR as a broadly safe and effective treatment option for patients with severe, symptomatic aortic stenosis (AS). Despite comprising a significant proportion of those requiring aortic valve replacement (AVR), patients with bicuspid AS were excluded from the pivotal trials due to several anatomic features such as non-circular annuli, heavier calcification, and coexisting aortopathy which were theorized to increase peri-procedural TAVR risk.^1^ Observational studies of highly-selected bicuspid AS patients undergoing TAVR have generally shown promising outcomes comparable to those with tricuspid AS.^2–5^ However, more direct observational comparisons of bicuspid AS undergoing TAVR and SAVR have reached mixed conclusions, likely due to variable degrees of confounding by indication and unmeasured patient characteristics.^6,7^ In the absence of randomized trial data comparing AVR strategies in patients with bicuspid AS, the optimal treatment approach in this subset of patients remains debated.^8,9^

On the basis of reassuring data from prospective registries of TAVR in bicuspid AS,^10–13^ the U.S. Food and Drug Administration (FDA) approved TAVR for low-risk patients regardless of aortic valve morphology (bicuspid or tricuspid) in 2019^14^ and subsequently removed a precaution in the labeling for TAVR use in patients with a congenital bicuspid aortic valve in 2020.^15,16^ Currently, U.S. guidelines support TAVR use in individuals age ≥ 65 years after shared decision-making that considers procedural risk, patient longevity, and valve durability.^17^ Though several studies have demonstrated that TAVR is expanding to younger populations^18–20^—where bicuspid AS is known to be more prevalent—the true utilization of TAVR in patients with bicuspid AS remains poorly understood. Present estimates suggest that 10% of all TAVRs are implanted in patients with bicuspid anatomy.^5,21^ However, among patients with bicuspid AS, the relative proportion of individuals receiving TAVR versus SAVR is unclear.

## Study Design

The TRanscatheter VErsus Surgical Aortic Valve ReplacemenT in Medicare Beneficiaries with Bicuspid Aortic Stenosis (TREAT-BICUSPID) study was designed to describe the trends of TAVR versus SAVR use in a population-wide analysis of Medicare beneficiaries ≥65 years of age with bicuspid AS, and to compare outcomes of TAVR and SAVR in patients with bicuspid AS at 1 year using an instrumental variable (IV) analysis. IV analysis is a quasi-experimental method derived from econometrics that leverages a source of random variation to account for both measured and unmeasured confounding and support causal inference in large observational datasets.^22^ We anticipated that TAVR use in bicuspid AS has been increasing following FDA low-risk approval, and given clinical equipoise surrounding the optimal management strategy for bicuspid AS, hospital-level preference in TAVR utilization could serve as a credible instrument that mimicked randomization within real-world data.^23^

## Methods

### Study Population

This retrospective population-based cohort study will be performed using Centers for Medicaid and Medicare Services (CMS) administrative data. The study has been granted a waiver of informed consent for retrospective data analysis by the Institutional Review Board of Beth Israel Deaconess Medical Center. All Medicare beneficiaries (Part A and Part B coverage, or managed care enrollment) age ≥65 years undergoing TAVR or SAVR between January 1, 2012, and December 31, 2022 with one year of claims data available before the procedure will be identified using *International Classification of Diseases, 9th Revision* (ICD-9) or *International Classification of Diseases, 10th Revision* (ICD-10) procedure codes. Individuals age <65 years and those with history of prior AVR are ineligible for this study. Similarly, patients undergoing percutaneous coronary intervention (PCI) for an acute coronary syndrome within 90 days prior to AVR date will be excluded, as this event commits a patient to an obligatory period of dual antiplatelet therapy and may affect subsequent preference for TAVR over SAVR.

Patients with bicuspid AS will be identified based on the presence of corresponding ICD-9 (746.3, 746.4) or ICD-10 (Q23.0, Q23.1) diagnosis codes. These codes have been previously validated and shown to have high specificity and positive predictive value for bicuspid aortic valves,^24^ and have been used previously to identify patients with bicuspid AS in other settings.^6,7,25,26^ Demographic information, including age, gender, race/ethnicity, will be obtained using CMS Master Beneficiary Summary File (MBSF) Base files. Baseline chronic conditions will be obtained through linkage of beneficiary data to the Chronic Conditions Warehouse (CCW), included as part of the Medicare Master Beneficiary Summary File. Additional baseline characteristics will be determined through a 1-year lookback period using Elixhauser Comorbidity Software or relevant diagnosis or procedure codes. Institutional characteristics will be derived from the American Hospital Association Annual Survey File. Procedure codes and diagnosis codes used in this study will be made available in the submitted manuscript.

### Outcomes

The primary endpoint of interest will be the composite of all-cause death, stroke, heart failure hospitalization, or aortic valve reintervention at 1 year. Secondary outcomes will include the individual components of the primary composite endpoint. Mortality will be derived from the MBSF Base files. Non-fatal outcomes will be identified using ICD-10 codes.

### Statistical Analysis

We will investigate trends in utilization by identifying the total number of patients with bicuspid AS undergoing AVR annually from 2012 to 2022. The number of patients undergoing (1) isolated TAVR, (2) TAVR with elective PCI in the 90 day period prior to AVR, (3) isolated SAVR, (4) SAVR with ascending aortic replacement (AAR), (5) SAVR with coronary artery bypass grafting (CABG), and (6) SAVR with other valve surgery will be calculated during each year of the study period. In the case of multiple surgical procedures performed at time of SAVR, a hierarchy of 1) AAR, 2) CABG, and 3) other valve surgery will be used to determine appropriate classification to reflect the strongest reasons as to why a patient with bicuspid AS would be treated surgically. In order to investigate the relative trends of TAVR and SAVR utilization among those who did not require additional cardiac surgery, we will examine trends in the subset of individuals who underwent isolated AVR. Trends will be evaluated in the overall study population and in prespecified subgroups defined by age and gender using the Cochran-Armitage trend test.

To identify predictors of utilization of TAVR over SAVR among the subset of patients being considered for isolated AVR, we will construct multivariable logistic regression models that will include patient demographics, baseline comorbidities, and hospital characteristics as candidate variables. The generalized estimating equations method will be used to account for within-hospital clustering due to similar AVR allocation strategy for patients treated at the same hospital. Annualized TAVR volume will be included in the model, calculated as the number of TAVRs performed at a hospital over the study period (inclusive of bicuspid and tricuspid patients), divided by the difference in the number of months between the first procedure and last procedure at that hospital, and then multiplied by 12. Models will be constructed for the entire study population and for younger patients aged 65-74.

Baseline characteristics of patients with bicuspid AS who received TAVR and SAVR over the study period will be presented as means ± standard deviation for continuous variables and counts with percentages for categorical variables. Standardized mean difference (SMD) will be used to compare treatment groups with a threshold of ≥0.1 to identify a clinically meaningful difference.

Given the critical importance of addressing both measured and unmeasured confounding when comparing outcomes between patients receiving TAVR or SAVR using observational data, we prespecified the use of an IV approach for the outcomes analysis. Whereas most customary statistical methods used in observational studies adjust only for measured characteristics and are therefore vulnerable to bias if treatment decisions are based on unmeasured characteristics, IV methods seek to leverage a source of naturally occurring variation in the data that balances both measured and unmeasured variables – thus more closely approximating experimental randomization and permitting causal effect estimates when several assumptions are met.^27–29^ Similar to true randomization, a valid instrument is causally related to treatment assignment, but unrelated to the outcome of interest. If certain key assumptions are met, a valid instrument recategorizes patients into treatment groups based on an increased likelihood of receiving the treatment assignment (i.e. TAVR), independent of baseline characteristics. Patients and outcomes are then compared according to an increased likelihood of receiving the treatment, rather than the actual treatment received.

We anticipate that though TAVR use in bicuspid AS has been broadly increasing following FDA low-risk approval, certain centers will have greater preference for one treatment strategy over the other during the study period, and hospital-level preference in TAVR utilization may serve as a credible instrument. Preference-based instruments in the medical literature have been widely described.^23,30–32^ Hospital-level preference for TAVR among bicuspid patients will be defined as the number of bicuspid TAVR procedures divided by the number of bicuspid AVR procedures performed at that site in the three previous calendar years, and will be recalculated for each hospital during each year of the study period. A three-year interval will be used to best capture a treating hospital’s preference for treatment strategy while minimizing the number of sites in which no bicuspid AS patients are treated with AVR in the calculation period. Accordingly, subjects who present to a hospital in which there were no bicuspid AS patients treated with AVR over the prior 3 years (denominator of hospital preference calculation = 0), and very low volume hospitals (<10 AVRs during any year of the study period) will be excluded from this analysis, as these hospitals may have patient populations and care patterns that may substantially differ from other hospitals in a manner that violates IV assumptions.

All Medicare beneficiaries age ≥65 years undergoing isolated TAVR or SAVR at the remaining hospitals between January 1, 2016, and December 31, 2021 will be eligible for the outcomes analysis. This time period has been selected in order to avoid inconsistencies between changes in ICD-9 and ICD-10 coding. All beneficiaries will be required to have 12 months of continuous enrollment both before and after the index AVR procedure to ascertain baseline comorbidities and clinical outcomes. Individuals with history of prior AVR will be excluded.

The IV outcomes analysis will be performed using a standard 2-stage least squares regression. In the first stage, the instrumental variable (hospital preference for bicuspid TAVR in prior three years) and all patient- and hospital-level characteristics are regressed on a binary outcome of receipt of TAVR or SAVR. Following this, coefficients obtained from the fully fitted first stage regression are used to calculate a predicted probability of receiving TAVR for every individual in the dataset. In the second stage, the clinical outcome of interest is regressed on the predicted probability of an individual receiving TAVR (endogenous variable) as estimated from the first stage model, adjusted for the same set of patient- and hospital-level characteristics used in the first stage. The absolute adjusted risk difference for the clinical outcome between patients treated with TAVR and SAVR can be derived from the coefficient of the endogenous variable (predicted probability of an individual receiving TAVR) from the second stage model. The procedure year will be included in both stages of the 2-stage least squares analysis to account for the contribution of time on both hospital preferences and outcomes during the study period.

The credibility of the IV approach is critically dependent on four necessary assumptions being met (**Table 1**). Only some of these assumptions can be objectively tested, whereas others must be reasoned. If these assumptions do not hold, an IV analysis is likely to be biased. First, the instrument should be relevant in that it should strongly predict the exposure of interest (“relevance”). This assumption can be empirically tested and will be assessed using the Cragg-Donald Wald F-test within the Stage 1 regression model to define the strength of the association between hospital-level preference for bicuspid TAVR and the likelihood that a patient with bicuspid AS presenting to that hospital receives TAVR, accounting for patient- and hospital-level characteristics. By rule, a partial F-statistic > 10 has been established as a threshold to identify a sufficiently strong instrument. Second, a valid instrument only affects outcomes through receipt of the treatment, and is otherwise unrelated to the outcome of interest except through the pathway of the treatment itself (“exclusion restriction”). This is a fundamentally untestable assumption. However, given the widely acknowledged clinical equipoise regarding best treatment strategy for bicuspid AS, it is plausible that hospitals with varying preference for bicuspid TAVR differ in their interpretation of existing evidence but are otherwise similar with no other mediators of clinical outcomes. Third, levels of the instrument must be as good as randomly assigned, such that any potential confounders are equally distributed among those who have an increased likelihood of receiving treatment (“exchangeability”). This is partially verifiable and will be evaluated through construction of baseline tables stratified across quintiles of the instrument. Lastly, the final assumption requires that there are no “defiers” in the dataset or patients with bicuspid AS who are less likely to receive TAVR if presenting to a hospital with a greater higher preference for bicuspid TAVR (“monotonicity”).

**Table 1:** Assumptions Required for Instrumental Variable Analysis

| Necessary IV Assumption | Definition | Interpretation within context of a hypothetical TAVR vs SAVR RCT | Interpretation within context of TREAT-BICUSPID observational study | Empirical tests available | Potential Violations and Consequences |
| --- | --- | --- | --- | --- | --- |
| Relevance | Instrument should predict the treatment of interest | Randomized treatment assignment (instrument) strongly predicts the treatment received (TAVR or SAVR). | Patients with bicuspid AS who present to a hospital with greater preference for bicuspid TAVR (instrument) should be more likely to receive TAVR (treatment), even after accounting for patient- and hospital- level characteristics | Cragg-Donald Wald F-test | Unlike an RCT, in which treatment assignment strongly commits a patient to TAVR or SAVR, the preference-based instrument provides a “nudge” towards a treatment strategy.<br>A weak instrument ($F < 10$) has poor correlation with the treatment and leads to biased and unreliable estimates. |
| Exclusion Restriction | Instrument only affects outcomes through the treatment | Random assignment to a treatment group only affects the outcome through the pathway of the actual treatment received.<br>This condition is best satisfied in double-blinded placebo controlled trials, where participants and study coordinators are unable to change their behavior based on knowledge of the treatment assignment. | Patients with bicuspid AS presenting to a hospital with greater preference for bicuspid TAVR have differential outcomes solely because of difference in receipt of TAVR. | None | For this assumption to hold, there must not be any other pathways in which a hospital with greater preference for bicuspid TAVR may influence clinical outcomes, aside from a patient with bicuspid AS being more likely to receive TAVR.<br>If hospitals with greater preference for bicuspid TAVR have consistently higher uptake of evidence-based medicine practices, or have greater resources for patient care, these could be alternative pathways aside from receipt of TAVR that may influence clinical outcomes.<br>If any of these alternative pathways to the outcome are present, then the effect estimate from IV analysis will be biased. |
| Exchangeability | Instrument is as good as randomly assigned | When performed effectively, randomization creates treatment groups that are otherwise similar aside from the treatment assignment received. | Patients with bicuspid AS that present to hospitals with greater preference for bicuspid TAVR must be similar to patients that present to hospitals with a lower preference for bicuspid TAVR. | Partially verifiable by looking at distribution of covariates across levels of the instrument.<br>Indirectly tested through falsification endpoints | If levels of the instrument are not as good as randomly assigned, and there exist imbalances in covariate distribution, there may be potential for confounding if those same covariates are associated with the outcome of interest. Residual confounding leads to biased estimates of causal effects.<br>Alternatively, imbalances in covariates that are not associated with the outcome may merely be observed differences. These do not always indicate the presence of residual confounding. |
| Monotonicity | Instrument consistently affects the treatment in a single direction | Randomization assigns patients to a treatment strategy, and all subjects are more likely to receive the assigned treatment after randomization.<br>There are no subjects who would seek out the alternative treatment once randomized. | Patients with bicuspid AS that present to a hospital with greater preference for bicuspid TAVR are more likely to receive TAVR in compliance with the recommendation of their local heart team.<br>There are no patients who would seek out the alternative treatment due to personal reasons or beliefs. | None | This assumption requires that the instrument only encourages or discourages the treatment in one direction, and is essential for valid estimation of the local average treatment effect. The causal effect isolated applies to those subjects in whom treatment was determined by the IV.<br>If there are “defiers” in the dataset, then the monotonicity assumption is violated and causal effect estimates can be biased. |

All diagnostics investigating the validity of the instrument will be performed prior to the TAVR versus SAVR outcomes analysis. To further evaluate for the presence of residual confounding in our primary outcomes analysis, we will prespecify the use of two falsification endpoints, hospitalization for pneumonia and hospitalization for urinary tract infection at 1 year, to be tested by the IV model. Falsification endpoints are endpoints that are not plausibly related in a causal manner to the treatment being evaluated, and thus serve as negative controls for an observational comparison.^33,34^ Pneumonia and hospitalization for urinary tract infection were chosen as they are unlikely to differ as a consequence of receiving TAVR or SAVR, but could differ if there remains unmeasured confounding in the IV analysis. Prespecified subgroup comparisons of TAVR versus SAVR will be performed by age (65-74, 75-84, ≥85) and sex (male/female). Absolute risk differences with Wald confidence intervals will be estimated, and two-sided p-values of <0.05 will be considered statistically significant. Data linkage and analyses will be performed using SAS 9.4 (SAS Institute, Cary, NC).

## Discussion

The TREAT-BICUSPID study has been designed to investigate the nationwide trends of TAVR versus SAVR use in individuals ≥65 years of age with bicuspid AS and attempt to provide evidence towards a key unanswered question regarding the optimal management strategy in these patients. As TAVR continues to expand to younger populations, where bicuspid AS is known to be more prevalent, there remains a need for high-quality evidence to resolve uncertainty and inform clinical practice. Recently, the NOTION-II study reported the 1-year outcomes of a low-risk population inclusive of bicuspid and tricuspid aortic valve anatomy randomized to TAVR or SAVR, and found potential harm from TAVR in the small subgroup of patients with bicuspid AS.^35^ Though there remains significant interest in a dedicated randomized trial of bicuspid AS patients, these trials remain under development, and results will not be available for many years.^9^

In the absence of randomized trials, observational studies are an important source of comparative data on different treatment modalities. However, most datasets are unlikely to capture the subtle but important prognostic variables that heart teams use to determine the best course of treatment for patients with bicuspid aortic valves, leaving such comparisons likely to be confounded. For this reason, our study prespecifies the use of an IV approach within existing observational data to exploit a real-word natural experiment that may better approximate the results of the types of randomized trials critically needed in this field. Our analysis requires that hospitals will differ in their use of TAVR in bicuspid AS patients, that patients with bicuspid AS are differentially assigned to a treatment strategy independent of measured characteristics, and that the hospital’s preference towards bicuspid TAVR is unrelated to other aspects of patient care that may influence clinical outcomes. If the requisite assumptions are met, this study will provide an unbiased comparison of aortic valve treatment strategies for the bicuspid AS population while large-scale, prospective trials are underway.

By design, the IV approach estimates a treatment effect for a “marginal” population, which is the subset of patients with bicuspid AS who would be treated with TAVR if they presented to a TAVR-leaning hospital, or SAVR if they presented to a SAVR-leaning hospital. The marginal population does not include the patients with bicuspid AS who would receive only one of the treatments under all circumstances no matter which hospital they presented, analogous to those who would be either be excluded from a clinical trial due to anatomy that is prohibitive for one treatment or the other, or, if included in the trial, crossover to receive the other treatment if randomized to the treatment they were ineligible to receive. Instead, the treatment effect isolated by this study design applies to the patients with bicuspid AS in whom either SAVR or TAVR could be considered. Defining the boundaries of this marginal population – based on Seivers subtypes, degree of leaflet calcification, presence of raphe, or extent of aortopathy – will be a key question for future studies.

## Limitations

The results of this analysis will need to be interpreted within the context of the study design. First, though Medicare data is useful for investigating trends over time, claims data lacks granularity on certain clinical features that may be pertinent to an outcomes analysis. Though we prespecified a statistical approach that could account for unmeasured confounding in this treatment comparison, the data does not permit stratification into levels of Society of Thoracic Surgeons (STS) surgical risk or analysis by transcatheter heart valve type (i.e. self-expanding versus balloon expandable valves), the latter of which may differ between hospitals. Second, the analysis proposed in this study investigates the effect of treatment decisions on clinical outcomes at 1 year. Patients with bicuspid AS undergoing TAVR may be at risk for worsened long-term durability due to paravalvular regurgitation, prosthesis undersizing, and valve thrombosis, all of which is not captured in this short-term study. Fortunately, use of claims data has shown promise for more complete tracking of outcomes after TAVR,^36^ and reanalysis of the cohort could be attempted in the future. Third, though the credibility of the IV will be tested rigorously, through diagnostic testing and falsification endpoints, some assumptions necessary for the IV analysis are fundamentally untestable. Lastly, despite providing an unbiased point estimate for treatment effect, IV approaches are associated with lower precision and may be underpowered if the true difference between groups is small.

## Conclusions

Patients with bicuspid AS were excluded from the pivotal TAVR trials, and clinical use of TAVR in bicuspid AS remains debated. Nevertheless, the FDA approved TAVR for low-risk patients regardless of aortic valve morphology in 2019, and multiple studies suggest that TAVR use in this population has been increasing. TREAT-BICUSPID is a retrospective, population-based cohort study of Medicare beneficiaries with bicuspid AS undergoing AVR from 2012 to 2022. The study will evaluate trends in relative use of TAVR and SAVR in bicuspid AS over time, and compare outcomes between treatment strategies using an IV approach. In the absence of randomized trial data, this study should provide novel and much needed evidence to guide management of bicuspid AS patients.

## Data Availability

All data produced in the present study are available upon reasonable request to the authors.

## Notes

### Competing Interest Statement

The authors have declared no competing interest.

### Funding Statement

This study did not receive any funding.

### Author Declarations

The Institutional Review Board of Beth Israel Deaconess Medical Center granted a waiver of informed consent for retrospective data analysis and gave ethical approval for this work.

